# CT-derived measurements of pulmonary blood volume in small vessels and need for oxygen in COVID-19 patients

**DOI:** 10.1101/2020.06.03.20121483

**Authors:** Wilfried De Backer, Muriel Lins, Wendel Dierckx, Jan Vandevenne, Jan De Backer, Ben R. Lavon, Maarten Lanclus, Karen Rijckaert, Irvin Kendall, Muhunthan Thillai

**Author notes:** Corresponding author: Prof. Dr. De Backer Wilfried, Faculty of Medicine, University of Antwerp. **Disclosure of interest** WD, JDB, BL, ML, IK, and KR are employees of FLUIDDA, a company that develops and markets part of the technology described in this paper. The other authors have no financial relationships with any organization or company that might have an interest in the submitted work and received no direct funding from FLUIDDA. No funding was received for this research. **Informed consent and patient details** The authors declare that this report does not contain any personal information that could lead to the identification of the patient(s). The authors declare that they obtained a written informed consent from the patients and/or volunteers included in the article. The authors also confirm that the personal details of the patients and/or volunteers have been removed.

## Abstract

In contrast to early reports of conventional acute respiratory distress syndrome (ARDS) as the underlying pathophysiology of hypoxemic respiratory failure observed in patients with severe COVID-19, more recent findings implicate direct involvement of the pulmonary vasculature in giving rise to these symptoms. In earlier research, we demonstrated that patients with COVID-19 showed markedly reduced pulmonary blood volumes in pulmonary vessels <5 mm^2^ in cross-sectional area visible on imaging (termed “BV5”), with attendant dilation of larger, more proximal vessels. Here, we present preliminary results in which reduced BV5 is shown to correlate significantly with increased need for supplemental oxygen and abnormal arterial blood gas measurements in hospitalized COVID-19 patients. We suggest a potential mechanistic link between observed clinical, pathological, and imaging findings, and outline how these may be helpful in clinical assessment as well as the development of novel therapies.

---

Throughout the COVID-19 pandemic, a portion of those affected have evolved towards acute hypoxic respiratory failure [1]. Initially this was hypothesized to result from acute lung injury leading to acute respiratory distress syndrome, with many COVID-19 patients meeting) the imaging portion of the Berlin criteria for ARDS [2, 3]. Treatment protocols for these patients were therefore designed based on the current guidelines for treatment of ARDS including mechanical ventilation with high levels of positive end-expiratory pressure (PEEP)[3]. A growing number of reports suggest that some patients present with relatively normal lung compliance but severe and refractory hypoxemia, which is inconsistent with the conventional understanding of ARDS, indicating that an alternate pathophysiology may be involved [4, 5, 6].

In previous research (Lins, et al), we used novel quantitative post-processing techniques of routine clinical chest CT scans to quantify the volume of blood contained within pulmonary blood vessels of a particular size, denoting these measurements “BVX”, where “X” indicates a range of vessel sizes in mm^2^ (BV5 is the volume of blood contained in vessels between 1.25 and 5 mm^2^ cross sectional area, BV5-10 between 5 and 10 mm^2^, and BV10 >10 mm^2^) [7]. We then expressed these quantities in terms of % total pulmonary blood volume for each individual patient. The results from scans of 108 healthy patients were compared to those from 103 patients with COVID-19. Patients with COVID-19 displayed a significant reduction in BV5 (expressed as % of total pulmonary blood volume) and an increase in BV5-10 and BV10, without significant differences in total pulmonary blood volume between both groups. These changes are equivalent to a redistribution of pulmonary blood away from the small pulmonary vessels and into larger vessels, a change consistent with increased pulmonary vascular resistance downstream. This increased resistance may be due to dysregulated vasoconstriction, thrombotic events in the microvasculature, or both. Any of these could contribute to the observed refractory hypoxemia by impairing gas exchange across the alveolar-capillary membrane. These data suggest a possible pathophysiological explanation for the impression of severe dead space ventilation reported by intensivists [6].

More recently, we performed further analysis of data from 51 hospitalized SARS-CoV2 positive patients at a single Belgian center, chosen based on the availability of paired clinical data alongside CT scans for most patients. All scans were provided under the oversight of the center’s local ethical committee with informed consent of patients. One patient was excluded due to highly anomalous arterial blood gases. 50 patients (30 male, 20 female, average age 61.98) were divided into groups based upon their need for oxygen at the time of CT scan acquisition, with a threshold of 2 L/m chosen a priori (<2 L/min O2 or > 2 L/min O2). BV5 values were compared between cohorts. Patients in the >2 L/min group had significantly lower BV5 values than patients in the < 2 L/min group. Furthermore, lower BV5 values were found to be significantly correlated with both increased alveolar-arterial oxygen gradient (AaDO2) and decreased PaO2 (Table 1). These new results give further credence to the notion that for many COVID-19 patients, the observed hypoxemia is a result of pathologic changes to pulmonary hemodynamics, leading to increased pulmonary vascular pressures and concomitant impairment of alveolar gas exchange. Early signs of treatment efficacy in pilot studies using heparin and inhaled nitric oxide further support the role of vascular dysfunction in the pathogenesis of COVID-19 [8, 9].

**Table 1.** Clinical Findings

|  | <2 L/m O2 (n = 32) | >2 L/m O2 (n= 18) | *p* | *R* |
| --- | --- | --- | --- | --- |
| BV5* (SD) | 46.89 (8.59) | 41.61 (6.95) | 0.023 | - |
| PaO2** vs. BV5 | - | - | 0.035 | 0.3 |
| AaDO2** vs. BV5 | - | - | 0.04 | -0.3 |

*\*BV5 as % total pulmonary blood volume*
*\*\*mmHG*

## Conclusions

Based on these preliminary findings we hypothesize that COVID-19 patients develop acute hypoxemic respiratory failure due, at least in part, to pathologic alterations in the pulmonary microvasculature and attendant impaired alveolar gas exchange. These changes may include occlusion of pulmonary blood vessels by thrombi, unusual vasoconstriction, and direct damage to the membranes across which gas exchange occur [2, 6, 10]. These vascular effects could inevitably lead to a degree of dead space ventilation. Therapeutic strategies in this phase of the disease should therefore focus on addressing these changes by way of vasodilators or anti-thrombotic strategies. Recruitment of distal airways with mechanical ventilation (with low levels of PEEP and high FIO2) may help to facilitate the transport of the inhaled oxygen towards the alveoli [5]. Those conducting clinical trials to assess the efficacy of these interventions should consider the use of small pulmonary blood vessel volumes as an endpoint.

## Data Availability

All patient data is available upon request from the corresponding author.

## References

1. Grasselli G, Zangrillo A, Zanella A, et al. Baseline Characteristics and Outcomes of 1591 Patients Infected With SARS-CoV-2 Admitted to ICUs of the Lombardy Region, Italy. JAMA. 2020;323(16):1574. doi:10.1001/jama.2020.5394

2. Ackermann M, Verleden SE, Kuehnel M, et al. Pulmonary Vascular Endothelialitis, Thrombosis, and Angiogenesis in Covid-19. N Engl J Med. Published online May 21, 2020:NEJMoa2015432. doi:10.1056/NEJMoa2015432

3. Meng L, Qiu H, Wan L, et al. Intubation and Ventilation amid the COVID-19 Outbreak: Wuhan’s Experience. Anesthesiology. 2020;132(6):1317–1332. doi:10.1097/ALN.0000000000003296

4. Gattinoni L, Coppola S, Cressoni M, Busana M, Rossi S, Chiumello D. Covid-19 Does Not Lead to a “Typical” Acute Respiratory Distress Syndrome. Am J Respir Crit Care Med. Published online March 30, 2020:prccm.202003-0817LE. doi:10.1164/rccm.202003-0817LE

5. Marini JJ, Gattinoni L. Management of COVID-19 Respiratory Distress. JAMA. Published online April 24, 2020. doi:10.1001/jama.2020.6825

6. Archer SL, Sharp WW, Weir EK. Differentiating COVID-19 Pneumonia from Acute Respiratory Distress Syndrome (ARDS) and High Altitude Pulmonary Edema (HAPE): Therapeutic Implications. Circulation. Published online May 5, 2020:CIRCULATIONAHA.120.047915. doi:10.1161/CIRCULATIONAHA.120.047915

7. Lins M, Vandevenne J, Thillai M, et al. Assessment of Small Pulmonary Blood Vessels in COVID-19 Patients Using HRCT. Radiology and Imaging; 2020. doi:10.1101/2020.05.22.20108084

8. Kobayashi J, Murata I. Nitric oxide inhalation as an interventional rescue therapy for COVID-19-induced acute respiratory distress syndrome. Ann Intensive Care. 2020;10(1):61. doi:10.1186/s13613-020-00681-9

9. Tang N, Bai H, Chen X, Gong J, Li D, Sun Z. Anticoagulant treatment is associated with decreased mortality in severe coronavirus disease 2019 patients with coagulopathy. J Thromb Haemost. 2020;18(5):1094–1099. doi:10.1111/jth.14817

10. Tan CW, Low JGH, Wong WH, Chua YY, Goh SL, Ng HJ. Critically ill COVID LJ19 infected patients exhibit increased clot waveform analysis parameters consistent with hypercoagulability. Am J Hematol. Published online May 4, 2020:pajh.25822. doi:10.1002/ajh.25822

